# Differential Lifespan Impacts on Veterans by War Exposure in the First World War

**DOI:** 10.1101/2022.09.29.22280532

**Authors:** Nick Wilson, Christine Clement, Jennifer A Summers, George Thomson, Glyn Harper

## Abstract

**Introduction:** There remains uncertainty around the impact of war on the lifespan of First World War (WW1) veterans.

**Methods:** We obtained lifespan data on a random sample of 857 war-exposed New Zealand WW1 veterans and compared this with lifespans of a non-war military cohort (n=1039). This comparison was possible as the non-war-cohort arrived in Europe too late to participate in the war, allowing a “natural experiment” that avoided the “healthy solider effect”.

**Results:** The lifespan comparisons indicated lower mean lifespan in the war-exposed veteran cohort compared to the non-war veteran cohort (69.7 vs 71.1 years; p=0.0405). This gap persisted (range: 0.8 to 1.1 years) but was no longer statistically significant when only considering the non-Māori ethnic grouping (nearly all European/Pākehā personnel), when excluding additional deaths in the immediate post-war period up to 31 December 1923, and when excluding participation in any other wars. Within the war-exposed cohort there were suggestive patterns of increasing lifespan with increasing occupational status and military rank (eg, 69.5, 70.0 and 70.7 mean years as group-level occupational status progressively increased). There were also stark differences in lifespan of 8.3 years between Māori (Indigenous) and non-Māori veterans (p=0.0083).

**Conclusions:** The pattern of reduced lifespan in war-exposed vs non-war-exposed veterans, was compatible with a smaller previous New Zealand study. There are a number of feasible avenues to further improve this type of work with existing data sources.

**Key messages:**

➢ There is still uncertainty around the impact of the First World War on premature death and lifespan in veterans internationally.
➢ This study of New Zealand veterans found lower mean lifespan in the war-exposed cohort compared to the non-war cohort (69.7 vs 71.1 years). This gap persisted, but was no longer statistically significant, when considering such factors as ethnicity of personnel and participation in any other wars.
➢ The pattern of reduced lifespan in war-exposed veterans was compatible with a smaller previous New Zealand study.
➢ There are a number of feasible avenues to further improve this type of work with existing data sources.

## INTRODUCTION

Some studies suggest that veterans of the First World War (WWI) had reduced lifespan, especially if they had adverse exposures and developed particular health conditions. A United Kingdom (UK) study found lower life expectancy in veterans awarded war pensions for either “disordered action of the heart (DAH)” or “neurasthenia/shellshock” when compared to a control group of gunshot wounded ex-servicemen.^1^ Similarly, a study of German veterans from WWI with head injuries found them to have increased mortality rates, relative to a control group of unwounded medal-winning veterans.^2^ In a United States (US) study, those veterans exposed to mustard gas had increased overall mortality rates relative to control groups of other veterans,^3^ including after further follow-up.^4^ Another study on mustard gas exposure among UK veterans, also found significantly higher mortality rates when compared to the male population of England and Wales.^5^ Finally, an Australian study which compared WW1 veterans with the general Australian male population reported relatively small lifespan impacts. These authors found that these veterans had “higher short-term mortality in 1919−1921, which results in lower life expectancy of 0.12 years, and higher longer-term mortality after 1921 which accounts for the remaining 0.53 years …”.^6^ This study also found that those more likely to die soon after the war “were older men (earlier birth year), taller men, and those from an unskilled working-class background”. For mortality after 1921, statistically significant risk factors in this Australian study were being discharged as medically unfit, being discharged as partially/totally permanently disabled, and being those discharged due to venereal disease.

Research on New Zealand WW1 veterans reported reduced lifespan when compared to a non-war cohort of military personnel who left this country for the war in late 1918, but who missed participating in the war.^7^ The use of this comparison group allowed for avoidance of the “healthy soldier effect” – a selection effect analogous to the “healthy worker effect” (as per a systematic review of this effect^8^ and also of the related “healthy warrior effect”^8 9^). Nevertheless, this New Zealand study was not particularly large and was potentially non-representative since the war-exposed cohort was those only leaving for the war in 1914 (with high exposure to the particularly adverse Gallipoli campaign). It also involved differences in terms of involvement of Māori (Indigenous) personnel who went to war from 1915 onwards, and it did not include conscripts who went to war from 1916 onwards. Hence we decided to repeat this type of study but with a larger random sample of veterans and which who covered the whole period of the war. With such a larger sample we also aimed to explore the potential impacts of other characteristics on lifespan including: ethnicity and socio-economic status. The more specific study aims are listed below.

1. To compare the lifespan of those war-exposed veterans who survived the war (from 1 January 1919) with an appropriate non-war comparison cohort. The latter reflected a “natural experiment” whereby the troopships carrying these personnel arrived too late in the war for them to see them participate.
2. To estimate differences in lifespan of veterans by ethnicity (Māori vs non-Māori).
3. To estimate differences in lifespan of veterans by pre-war occupation (as a proxy of socio-economic status).

To provide context to the New Zealand role in WWI, there were an estimated 98,950 military personnel who served outside of the country, and 7036 who served on home territory in the New Zealand Expeditionary Force (NZEF).^10^ The former personnel mainly fought in Europe on the Western Front, but also were part of campaigns in the Middle East (Gallipoli and Palestine).^11^ An estimated 16.6% died during the war^12^ at least to the period to Armistice day in 1918, with others dying subsequently raising the total to an estimated 18,311 by the end of 1923 (18.2% of participants). The latter time period was that used in the official New Zealand *Roll of Honour*.^10^ The official number of personnel wounded or suffering illness was 41,316 (with these involving removal from the front-line for medical treatment).^10^ These mortality and morbidity burdens do not include the more than 12,000 New Zealanders who served separately with other military forces during this war, with just over 1400 of these dying in combat.^13^

## METHODS

### Selection of war-exposed veterans

In February 2021, we downloaded data on all WW1 participants from Auckland War Memorial Museum’s Cenotaph website database.^14^ This was 104,993 individual names, which is 99.1% of war participants according to an official estimate of 105,986.^10^ Then within this dataset we randomly selected a 5% sample of personnel (using the random number generator in Microsoft Excel). From this sample we then excluded:

- Non-veterans (defined as those dying in the war and subsequent weeks up to 31 December 1918), with this group being 18.2% of the sample.
- Female participants (who were nurses and volunteers), with this being 0.47% of the sample.
- Pasifika military personnel who came from South Pacific Islands to join the NZEF, with this group being 0.50% of the sample. This grouping was excluded given the focus of the study was on veterans from New Zealand and because there is large heterogeneity in backgrounds for Pasifika populations (demography and culture). These Pasifika individuals were identified based on the home address of themselves or next of kin (using word searches in the dataset for the names: Niue, Cook Islands/Rarotonga, Samoa, Fiji, and Tokelau).

But then given financial resourcing constraints on the study, we then focused on just 1000 of the remaining group (as determined by the ordering of their assigned random numbers). Further exclusions from this group of 1000 once archival military records were examined in more detail, included 113 individuals (11.3%). These were on the grounds of:

- Being a non-veteran (as per the definition above) on closer examination of the records.
- Not seeing active war service overseas (eg, not actually leaving New Zealand, deserting on route or being deemed medically unfit prior to war exposure)
- Only serving in Samoa (since this theatre of the war involved no conflict with Germany).
- They were actually part of a non-New Zealand military force (eg, the Australian Imperial Force (AIF), or the Royal Navy of the UK).

We acknowledge that the term “war-exposed” is a simplification for this selected group as some individuals may have been in support roles that were far from the front-line. But such case-by-case determinations would usually only be possible after very detailed analysis of each military file, and even then sometimes not.

### Selection of non-war veterans

To allow for a non-war comparison cohort of veterans, we selected all the male military personnel on two New Zealand troopships which departed near the end of the war and did not participate in it. These troopships were the *Athenic* (His Majesty’s NZ Transport [HMNZT] 106) which departed on 13 June 1918 with 793 military personnel and the *Ulimaroa* (HMNZT 108) which departed on 27 July 1918 with 1010 military personnel. The lifespan details of these personnel were described in a previous study^15^ in which comparisons were made with another troopship that suffered from a severe onboard outbreak of pandemic influenza (ie, the troopship *Tahiti* [HMNZT 107]). Although there were no statistically significant lifespan differences found by exposure to pandemic influenza, just in case some small such impact was missed, we decided to focus on using the data from the personnel on the *Athenic* and *Ulimaroa* (and not the *Tahiti*).

### Lifespan data

We collected data on birth dates from the online military files.^16^ Date of death was also sometimes in these files but otherwise we used a range of genealogical sources. These include the Births Deaths and Marriages database^17^ (which should contain records of all New Zealand-based deaths in these cohorts), and where there was an exact match between the name, and age and year of death with the date of birth data from the military file. Other genealogical sources included online cemetery records and the Cenotaph database. In some cases, only the birth year or year of death could be identified, so we used the mid-year point of such years (eg, 1 July 1890) in the analyses to calculate the age at death. Furthermore, when the reported age at death was recorded in years only, we used a mid-point value, ie, age “70 years” was adjusted to “70.5 years”.

In the various lifespan analyses we made allowance for ethnicity of personnel, participation in other wars and death early in the post-war period (1919 to 1923).

### Coding ethnicity, class, and military rank

Māori ethnicity was assumed if: (i) the person had any name in te reo Māori; or (ii) the person served in a Māori Contingent/Pioneer Battalion plus had either: (a) a next of kin with a name in te reo Māori; or (ii) the person had a stated iwi affiliation on the Cenotaph website. We acknowledge however that this approach will have missed some personnel who self-identified as Māori as some participated outside of the Pioneer Battalion and some used only European/Pākehā names. But identifying these personnel was out of scope for this study as it would require very extensive genealogical research. Participation in the Pioneer Battalion alone was not considered enough to imply Māori ethnicity as this Battalion did not become a fully Māori one until 1 September 1917.

Occupational class was classified in the same way as two previous studies^15 18^ by being based on the stated occupation at enlistment in online military files. A table in the Supplementary Material of one of these studies^18^ gives the full list of occupations and their associated classifications.

For military rank we considered both the first rank on enlistment and highest rank achieved as listed in the military files. These were analysed according to three groupings: (i) Commissioned Officers; (ii) all Non-Commissioned personnel excluding the lowest rank; (iii) the lowest rank (eg, gunner, trooper, sapper, signaller, and private).

### Data analysis

Most analyses involved comparing mean lifespan using an ANOVA, but where the data were not normally distributed a non-parametric test was used ie, Kruskal-Wallis test for two groups. A p-value of <0.05 was considered statistically significant.

### Ethics statement

Ethical approval was provided through the University of Otago Human Ethics Committee process (Category B Approval, D22/030).

## RESULTS

After a range of exclusions made to the randomly-selected sample (Table 1), the sub-sample of 1000 personnel had 887 deemed eligible for inclusion. Of these, we were able to locate birth and death data to estimate age at death for 857 (96.6%). This group was nearly entirely non-Māori personnel, though 2.7% were classified as being Māori. A majority (60.4%) were from the lowest three occupational class groupings (on a nine-point scale, Table 1). Similarly, a majority (86.8%) were in the lowest military ranks on enlistment (eg, gunner, trooper, sapper, signaller, and private).

**Table 1:** Description of the random sample of New Zealand military personnel participating in the First World War who were defined as war-exposed and who were alive from 1 January 1919

| Sample description | Number | % |
| --- | --- | --- |
| **Full list of NZEF participants** in the Cenotaph database supplied by Auckland War Memorial Museum in February 2021 | 104,993 | 100% |
| **Random** 5% sample of the above | 5250 | 5.0% (of total) |
| Removal of non-New Zealand based military personnel who came from various **Pacific Islands** (see *Methods*) | 26 | 0.50%<br>(26/5250) |
| Removal of **female participants**. This was based on first names and then also checking that the occupation was either “nurse” or “volunteer” (the only known occupations for female personnel in the NZEF). | 25 | 0.47%<br>(25/5250) |
| Removal of **those dying in the war** and subsequent weeks up to 31 December 1918 | 953 | 18.2%<br>(953/5250) |
| **More focused sample of 1000 active service personnel (a random subset of the above sample)** |  |  |
| Further exclusions based on more detail examination of the military records: (i) Did not participate in the war overseas (eg, arrived in France after the war ended), n=45 (39.8%); (ii) Served in a foreign military such as the Australian or UK forces, n=31 (27.4%); (iii) Declared medically unfit before participating in the war (eg, a disability that was initially missed when recruited was identified or one arose before going to the front-line), n=22 (19.5%); (iv) only served in Samoa where there was no conflict with Germany, n=6, (5.3%); (v) Deserted before participating in the war, n=4, (3.5%); (vi) Died in late 1918 (see above), n=2 (1.8%); (vii) Were from a Pacific Island (see above), n=2 (1.8%); (viii) Were female, n=1, 0.9%) | 113 | 11.3%<br>(113/1000) |
| Age at death *could not* be ascertained (eg, potentially these individuals moved to a country after WW1 with non-accessible death records; or they changed their name) | 30 | 3.4%<br>(30/887) |
| Age at death *could* be ascertained | 857 | 96.6%<br>(857/887) |
| **Characteristics of the n=857 with age at death data** |  |  |
| **Ethnicity** |  |  |
| – Māori (see *Methods* for definitions) | 23 | 2.7% |
| – Non-Māori ethnicity (nearly all European/Pākehā) | 834 | 97.3% |
| **Occupational class\*** (based on occupation at enlistment) |  |  |
| – Highest 3 groupings (ie, least deprived) | 51 | 6.0% |
| – Middle 3 groupings | 288 | 33.6% |
| – Lowest 3 groupings (ie, more deprived) | 517 | 60.4% |
| **War-related aspects** |  |  |
| Also participated in the South African War (1899-1902) | 8 | 0.9% |
| Also participated in WW2 (1939-1945) | 62 | 7.2% |
| Prisoner-of-war in WW1 (1914-1918) | 5 | 0.6% |
| **Military rank on enlistment\*** |  |  |
| – Commissioned Officers (ie, from officer cadet to field marshal) | 22 | 2.6% |
| – All Non-Commissioned personnel (excluding the lowest rank) (ie, from lance corporal to warrant officer class one) | 91 | 10.6% |
| – The lowest rank (eg, gunner, trooper, sapper, signaller, private) | 743 | 86.8% |
| **Military rank – highest achieved\*** |  |  |
| – Commissioned Officers | 44 | 5.1% |
| – All Non-Commissioned personnel (excluding the lowest rank) | 166 | 19.4% |
| – The lowest rank (eg, private) | 646 | 75.5% |
| **Military rank – changes during the war out of 18 possible levels\*** |  |  |
| – No change | 658 | 76.9% |
| – Increased | 164 | 19.2% |
| – Decreased | 34 | 4.0% |
| – Mean change in rank | 0.42 | NA |
| – Range of changes in rank | -3 to +8 | NA |
| **War-related death reported** from 1 January 1919 onwards (eg, died from wounds, accidental death as occupation force, infectious disease related to war service, or killed in WW2)* |  |  |
| – WW1 | 9 | 1.1% |
| – WW2 | 4 | 0.5% |

\* Missing data for 1 individual (military file missing from archives), with % calculated out of n=856.

Nevertheless, by the end of the war 19.2% had increased their military rank, an average of a 0.42 step increase out of 18 levels of military rank. A small proportion of these 857 personnel (0.9%) had previously participated in the South African War and 7.2% subsequently went on to participate in the Second World War (WW2).

Despite being alive on 1 January 1919, this group continued to experience WW1-related deaths (n=9 deaths, 1.1%, Table 1). These were from the impact of wounds, accidental death while in an occupation force, and infectious disease related to war service. Some also died as part of service in WW2 (n=4, 0.5%).

### Results of the lifespan analyses – war-exposed cohort

For the group of 857 personnel with lifespan data, there were no statistically significant differences in lifespan by birth decade/period, occupational status on enlistment or by military rank (on enlistment, highest rank achieved and progression in rank) (Table 2). Nevertheless, there were suggestive patterns of increasing mean/median lifespan with increasing occupational status and military rank (eg, 69.5, 70.0 and 70.7 mean years as group-level occupational status progressively increased). There were however, stark differences in veteran lifespan by ethnicity of 8.3 years (means of 61.7 for Māori and 70.0 for non-Māori; p=0.0083). A war-related death from 1919 onward (either as a consequence of WW1 or participation in WW2) was also related to markedly lower lifespan (40.0 years vs 70.2 years; p<0.00001, Table 2). Nevertheless, overall participation in other wars (South African War or WW2) was not associated with reduced lifespan relative to those who only participated in WW1.

**Table 2:**
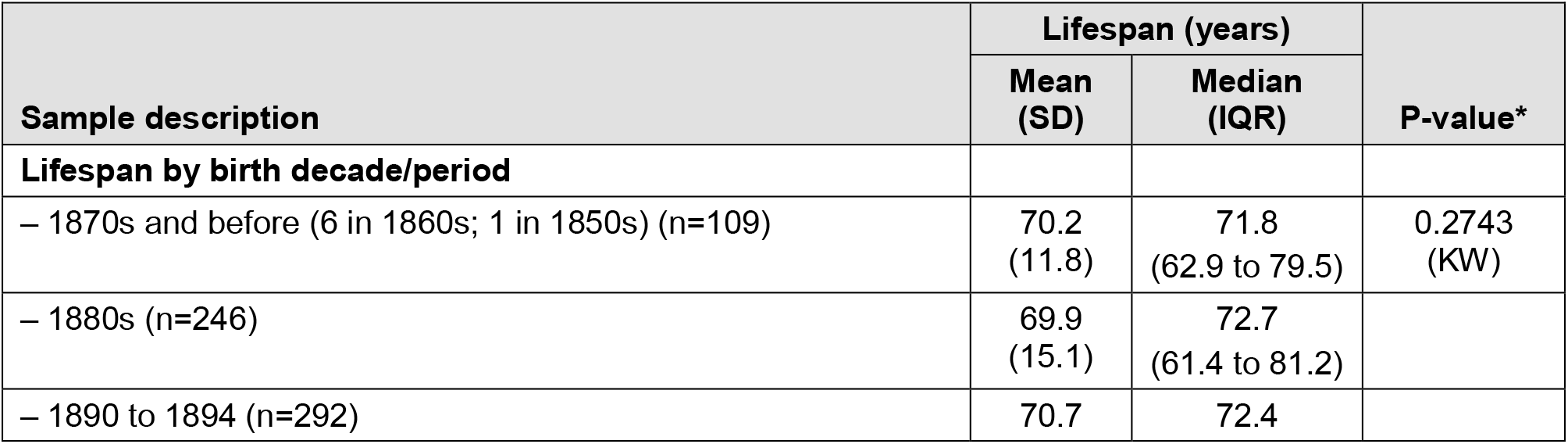
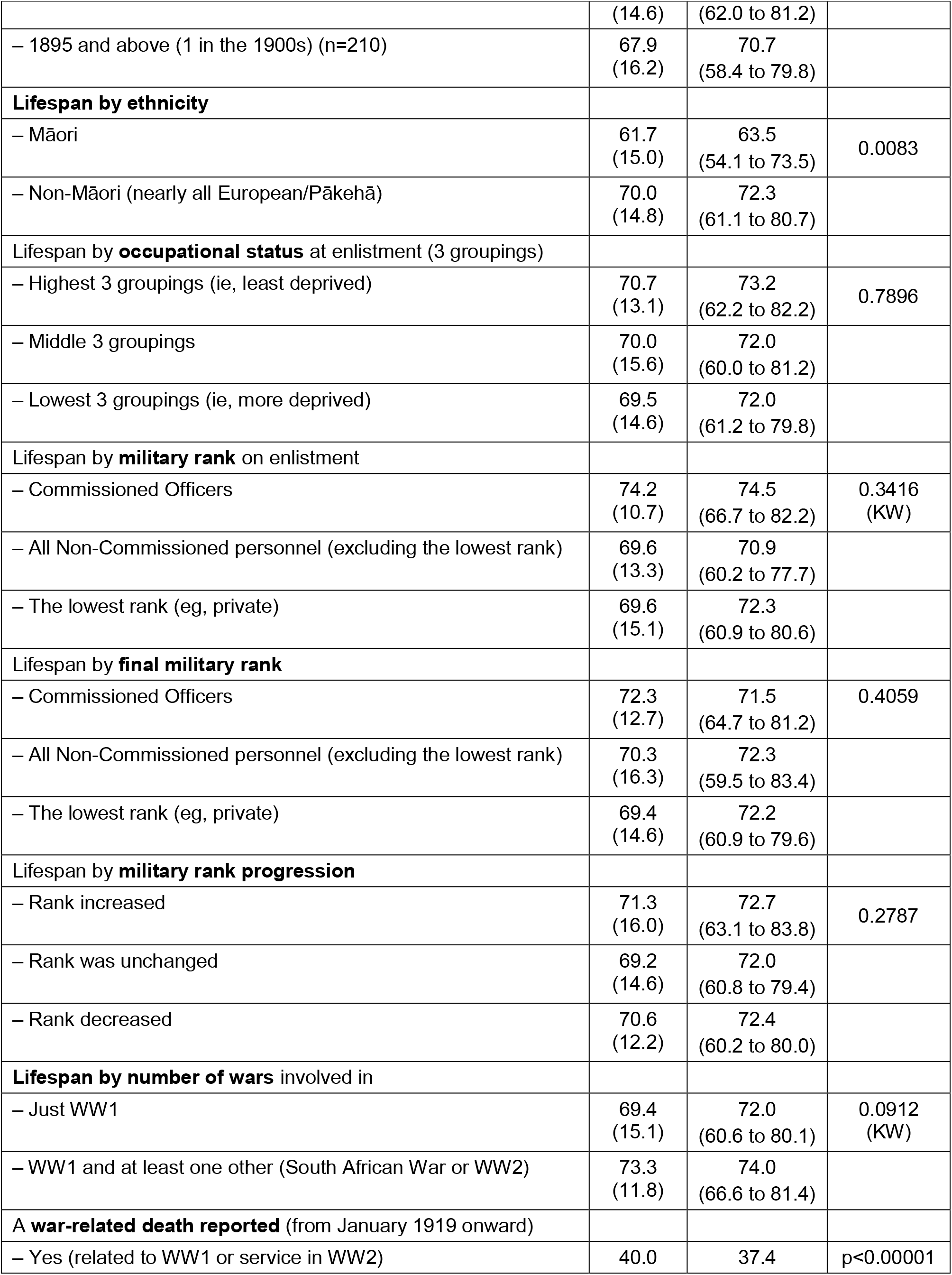
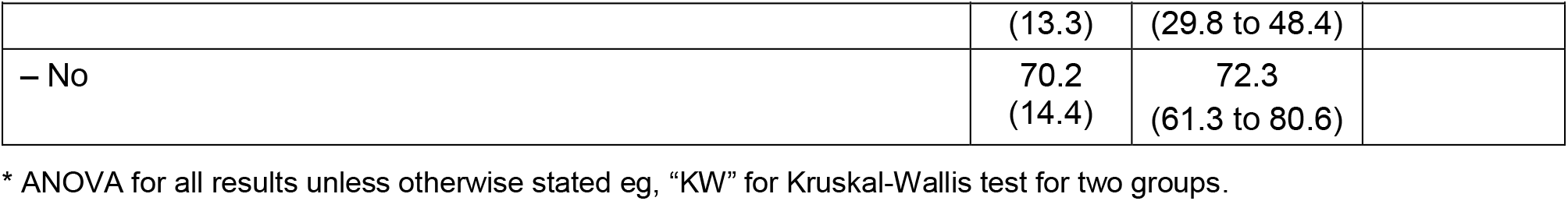
Results of the lifespan analyses of the New Zealand military personnel participating in the First World War who were defined as war-exposed veterans and who were alive from 1 January 1919 (for a random sample of 857)

### Results of the lifespan analyses – war-exposed vs non-war cohorts

The socio-demographic characteristics of the war-exposed and non-war cohorts are compared in Table 3. They had identical median birth years but the mean year of birth was slightly higher (1.0 years) in the non-war group (p=0.0030). Another difference of note was the lower proportion of Māori personnel in the non-war cohort compared to the war-exposed cohort (0.2% vs 2.7%; p<0.00001).

**Table 3:**
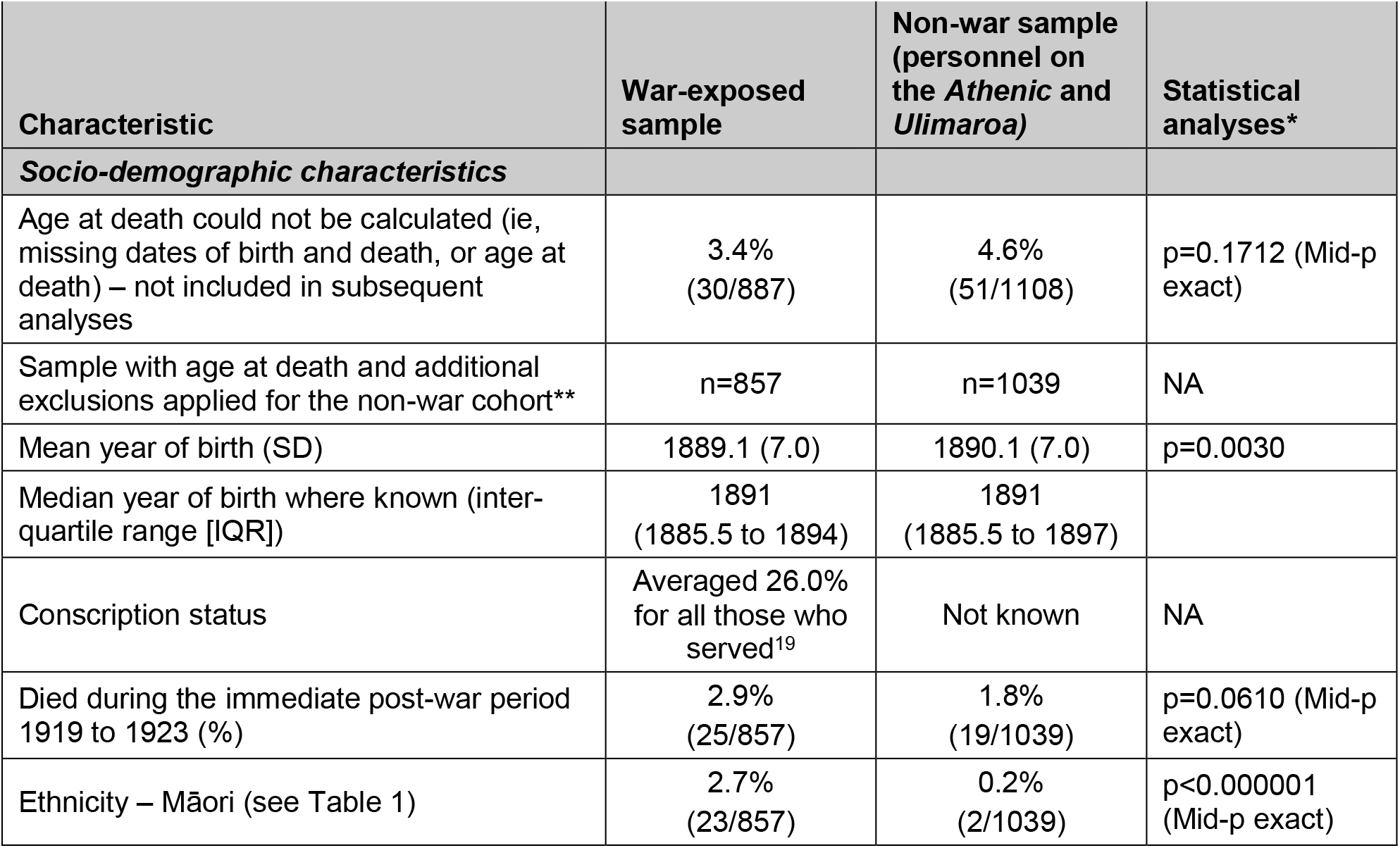
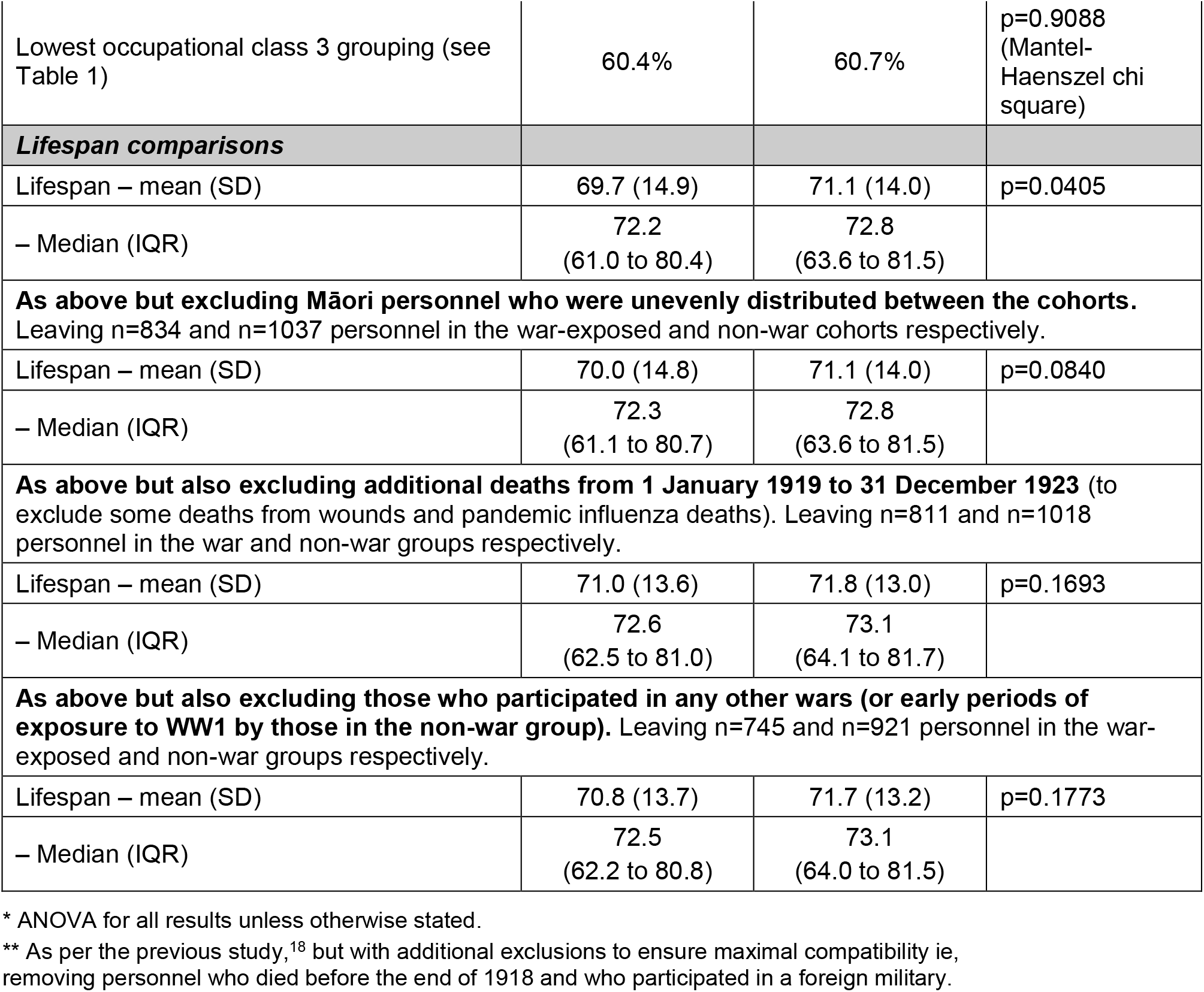
Comparison of characteristics and lifespan between the war-exposed and non-war veterans (cohorts of New Zealand military personnel alive on 1 January 1919)

| Characteristic | War-exposed sample | Non-war sample (personnel on the *Athenic* and *Ulimaroa*) | Statistical analyses* |
| --- | --- | --- | --- |
| ***Socio-demographic characteristics*** |  |  |  |
| Age at death could not be calculated (ie, missing dates of birth and death, or age at death) – not included in subsequent analyses | 3.4%<br>(30/887) | 4.6%<br>(51/1108) | $p=0.1712$ (Mid-p exact) |
| Sample with age at death and additional exclusions applied for the non-war cohort** | n=857 | n=1039 | NA |
| Mean year of birth (SD) | 1889.1 (7.0) | 1890.1 (7.0) | $p=0.0030$ |
| Median year of birth where known (inter-quartile range [IQR]) | 1891<br>(1885.5 to 1894) | 1891<br>(1885.5 to 1897) |  |
| Conscription status | Averaged 26.0% for all those who served<sup>19</sup> | Not known | NA |
| Died during the immediate post-war period 1919 to 1923 (%) | 2.9%<br>(25/857) | 1.8%<br>(19/1039) | $p=0.0610$ (Mid-p exact) |
| Ethnicity – Māori (see Table 1) | 2.7%<br>(23/857) | 0.2%<br>(2/1039) | $p<0.000001$ (Mid-p exact) |
| Lowest occupational class 3 grouping (see Table 1) | 60.4% | 60.7% | p=0.9088 (Mantel-Haenszel chi square) |
| **Lifespan comparisons** |  |  |  |
| Lifespan – mean (SD) | 69.7 (14.9) | 71.1 (14.0) | p=0.0405 |
| – Median (IQR) | 72.2<br>(61.0 to 80.4) | 72.8<br>(63.6 to 81.5) |  |
| **As above but excluding Māori personnel who were unevenly distributed between the cohorts. Leaving n=834 and n=1037 personnel in the war-exposed and non-war cohorts respectively.** |  |  |  |
| Lifespan – mean (SD) | 70.0 (14.8) | 71.1 (14.0) | p=0.0840 |
| – Median (IQR) | 72.3<br>(61.1 to 80.7) | 72.8<br>(63.6 to 81.5) |  |
| **As above but also excluding additional deaths from 1 January 1919 to 31 December 1923 (to exclude some deaths from wounds and pandemic influenza deaths). Leaving n=811 and n=1018 personnel in the war and non-war groups respectively.** |  |  |  |
| Lifespan – mean (SD) | 71.0 (13.6) | 71.8 (13.0) | p=0.1693 |
| – Median (IQR) | 72.6<br>(62.5 to 81.0) | 73.1<br>(64.1 to 81.7) |  |
| **As above but also excluding those who participated in any other wars (or early periods of exposure to WW1 by those in the non-war group). Leaving n=745 and n=921 personnel in the war-exposed and non-war groups respectively.** |  |  |  |
| Lifespan – mean (SD) | 70.8 (13.7) | 71.7 (13.2) | p=0.1773 |
| – Median (IQR) | 72.5<br>(62.2 to 80.8) | 73.1<br>(64.0 to 81.5) |  |

\* ANOVA for all results unless otherwise stated.
\*\* As per the previous study,<sup>18</sup> but with additional exclusions to ensure maximal compatibility ie, removing personnel who died before the end of 1918 and who participated in a foreign military.

The lifespan comparisons indicated a significantly lower mean lifespan (1.4 years) in the war-exposed cohort compared to the non-war cohort (69.7 vs 71.1 years; p=0.0405; Table 3). This gap persisted (range: 0.8 to 1.1 years) but was no longer statistically significant when only considering the non-Māori ethnic grouping (nearly all European/Pākehā personnel), when excluding additional deaths in the immediate post-war period up to 31 December 1923, and when excluding participation in any other wars.

## DISCUSSION

### Main findings and interpretation

This analysis found relatively small lifespan difference (1.4 years) between the war-exposed cohort compared to the non-war cohort (69.7 vs 71.1 years; p=0.0405). Such a lifespan gap persisted, but was no longer at a statistically significant level in other analyses (eg, when only considering the other (non-Māori) ethnic grouping and excluding those who participated in other wars). The findings are relatively similar to a previous New Zealand study that found a 1.7 year lower median lifespan for war-exposed veterans, albeit when considering a smaller cohort of those who left for the war in 1914.^7^ This finding is also compatible with the international literature for WW1 (see *Introduction*) – although all the comparison groups used in these studies are not as appropriate as these New Zealand studies. Other New Zealand work for other wars has also suggested reduced lifespan of war-exposed veterans (one WW2 study^20^), but not in another WW2 study^21^ and not in a South African War study.^22^

The large differences in veteran lifespan by ethnicity (8.3 years less for Māori vs non-Māori personnel) were unsurprising and reflect very long-term health disparities by ethnicity.^23^ Reducing such health inequalities is an ongoing goal for the current New Zealand health sector and government.

The pattern of increasing lifespan with higher occupational class (albeit not at a statistically significant level) was also an unsurprising finding. This pattern has previously been described in another study of New Zealand military personnel in WW1.^18^ Perhaps more surprising was that participation in other wars (South African War or WW2) was not associated with reduced lifespan relative to those who only participated in WW1. This could reflect a mix of self-selection effects and military selection procedures whereby those with health problems were less likely to participate in more than one war. Another reason might have been that those going on to participate in WW2, did so with higher military ranks or in training roles and were therefore less likely to be subjected to hardship/injuries/illness on the front-line.

### Study strengths and limitations

A strength of this study was its relatively large size and its sampling from all New Zealand military participants of WWI (compared to the previous work with its focus on only those leaving in 1914^7^). It was able to utilise a “natural experiment” involving relatively comparable cohorts (war-exposed vs non-war) which had been through a similar selection process into the military as so avoid the “healthy solider effect”.^8^ This study is therefore probably one of the most detailed to date of lifespan of military personnel arising from participation in WW1 – alongside an Australian study which made lifespan comparisons with the general male population.^6^

Nevertheless, a number of limitations with our study need to be considered. In particular, the cohorts were not perfectly comparable, as shown in Table 2 (ie, identical median birth years but a 1.0 year difference in mean birth year). Other group-level differences of possible relevance included health status on selection into the military (including height, weight and medical conditions; for which we did not extract data), and conscription status (which is not recorded in the military files).

Also, post-war experiences by cohort probably differed as the non-war cohort may have returned to New Zealand from war later in 1919 than the war-exposed cohort, and therefore had poorer access to the limited employment opportunities available.

### Further research possibilities

The legacy impacts of war to both veterans and civilians are important to understand – as warfare is a continuing problem in the modern era. Therefore this type of study could be expanded if it was possible to fund genealogical, health and military researchers to conduct work that reduced the relevance of the study limitations detailed above. For example, more detailed analysis of individual military files and other archival records could determine such issues as biometric data on selection into the military, proximity to front-line combat in various theatres of war, and exposure to unemployment on return to New Zealand. But some risk factor data might never be obtainable eg, smoking status does not seem to be available in any records. In New Zealand it is also possible to obtain the death certificates on all WW1 personnel which could be used to establish causes of deaths that may be conflict-related (eg, mustard gas exposure causes long-term respiratory disease;^24^ suicide related to post-traumatic stress disorder; and death after surgery on wounds continued after the war). However, researchers still have to purchase such death certificates and so such a study would require reasonable resourcing. All such research would ideally be funded by Veterans’ Affairs New Zealand or the New Zealand Defence Force. Alternatively, the Ministry of Culture and Heritage could support work in this major domain of New Zealand’s history.

## CONCLUSIONS

This study found lower mean lifespan in the war-exposed cohort compared to the non-war-exposed cohort (69.7 vs 71.1 years). This gap persisted, but was no longer statistically significant, when considering such factors as ethnicity of personnel and participation in any other wars. Nevertheless, the pattern of reduced lifespan in war-exposed veterans was compatible with a smaller previous New Zealand study and an Australian study. There are a number of feasible avenues to further improve this type of work with existing data sources.

## Data Availability

All data produced in the present study are available upon reasonable request to the authors

## Contributors

NW conceived the idea and managed the project. GH obtained the research funding support. CC and NW gathered and processed the appropriate data. NW conducted the analyses and wrote the first draft. All authors contributed to interpretation and revising multiple drafts of the manuscript.

## Acknowledgements

We thank the Auckland War Memorial Museum and the Cenotaph Database Team for collating the data on NZEF personnel and making the data available free of charge. We thank Massey University for research funding that was provided to one co-author (CC).

## Funding

Massey University research funding was provided to one co-author (CC).

### Competing interests

None declared.

### Patient consent for publication

Not applicable.

### Ethics approval

Ethical approval was provided by the University of Otago Human Ethics Committee (Category B Approval, reference number D22/030).

### Provenance and peer review

Not commissioned; externally peer reviewed.

### Data availability statement

Data are available upon reasonable request.

## Notes

### Competing Interest Statement

The authors have declared no competing interest.

## References

1. Jones E, Vermaas RH, Beech C, Palmer I, Hyams K, Wessely S. Mortality and postcombat disorders: U.K. veterans of the Boer War and World War I. Mil Med 2003;168:414–8.

2. Weiss GH, Caveness WF, Einsiedel-Lechtape H, McNeel ML. Life expectancy and causes of death in a group of head-injured veterans of World War I. Arch Neurol 1982;39:741–3.

3. Beebe GW. Lung cancer in World War I veterans: possible relation to mustard-gas injury and 1918 influenza epidemic. J Natl Cancer Inst 1960;25:1231–52.

4. Norman JE, Jr. Lung cancer mortality in World War I veterans with mustard-gas injury: 1919-1965. J Natl Cancer Inst 1975;54:311–7.

5. Case RA, Lea AJ. Mustard gas poisoning, chronic bronchitis, and lung cancer; an investigation into the possibility that poisoning by mustard gas in the 1914-18 war might be a factor in the production of neoplasia. Br J Prev Soc Med 1955;9:62–72.

6. McCalman J, Kippen R, McMeeken J, Hopper J, Reade M. Early Results From the ‘Diggers to Veterans’ Longitudinal Study of Australian Men who Served in the First World War. Short-and Long-Term Mortality of Early Enlisters. Historical Life Course Studies 2019;8:52–72.

7. Wilson N, Clement C, Summers JA, Bannister J, Harper G. Mortality of first world war military personnel: comparison of two military cohorts. BMJ 2014;349:g7168.

8. McLaughlin R, Nielsen L, Waller M. An evaluation of the effect of military service on mortality: quantifying the healthy soldier effect. Ann Epidemiol 2008;18:928–36.

9. Wilson J, Jones M, Fear NT, Hull L, Hotopf M, Wessely S, Rona RJ. Is previous psychological health associated with the likelihood of Iraq War deployment? An investigation of the “healthy warrior effect”. Am J Epidemiol 2009;169:1362–9.

10. Ministry for Culture and Heritage. First World War by the numbers. Ministry for Culture and Heritage (Updated 25 February 2020). URL: https://nzhistory.govt.nz/war/first-world-war-by-numbers.

11. Crawford J, McGibbon IE. New Zealand’s Great War: New Zealand, the Allies, and the First World War. Auckland: Exisle Publishing Ltd, 2007.

12. Wilson N, Summers JA, Baker MG, Thomson G, Harper G. Fatal injury epidemiology among the New Zealand military forces in the First World War. N Z Med J 2013;126:13–25.

13. Harper G, Clement C, Johns R. For King and Other Countries. Auckland: Massey University Press, 2019.

14. Auckland War Memorial Museum. Cenotaph database. (Accessed July 2020). https://www.aucklandmuseum.com/war-memorial/online-cenotaph.

15. Wilson N, Boyd M, Nisa S, Clement C, Baker MG. Did exposure to a severe outbreak of pandemic influenza in 1918 impact on long-term survival? Epidemiol Infect 2016;144:3166–69.

16. New Zealand Government. Archway. Archives New Zealand. https://www.archway.archives.govt.nz

17. Department of Internal Affairs. Births, Deaths & Marriages Online. https://www.bdmhistoricalrecords.dia.govt.nz/search/.

18. Wilson N, Clement C, Boyd M, Teng A, Woodward A, Blakely T. The long history of health inequality in New Zealand: occupational class and lifespan in the late 1800s and early 1900s. Aust N Z J Public Health 2018;42:175–79.

19. Census and Statistics Office. The New Zealand Official Year-Book, 1919. Wellington: Census and Statistics Office, 1920. https://www3.stats.govt.nz/New_Zealand_Official_Yearbooks/1919/NZOYB_1919.html.

20. Wilson N, Harper G. Lifespan of New Zealand Second World War veterans from one large cemetery: the case for a national-level study. N Z Med J 2019;132:96–98.

21. Wilson N, Clement C, Summers J, Thomson G, Harper G. Impact of war on veteran life span: natural experiment involving combat versus non-combat exposed military personnel. BMJ Mil Health 2021.

22. Wilson N, Clement C, Thomson G, Harper G. Health impacts for New Zealand military personnel from the South African War of 1899-1902. N Z Med J 2021;134:22–43.

23. Woodward A, Blakely T. The Healthy Country? A History of Life and Death in New Zealand. Auckland: Auckland University Press, 2014.

24. Tang FR, Loke WK. Sulfur mustard and respiratory diseases. Crit Rev Toxicol 2012;42:688–702.

